# From voice to ink (VINK): Development and assessment of an automated, free-of-charge transcription tool

**DOI:** 10.1101/2023.05.04.23289518

**Authors:** Hannah Tolle, Maria del Mar Castro, Jonas Wachinger, Agrin Zauyani Putri, Dominic Kempf, Claudia M. Denkinger, Shannon A. McMahon

## Abstract

Verbatim transcription of qualitative data is a cornerstone of analytic quality and rigor, yet the time and energy required for such transcription can drain resources, delay analysis and hinder the timely dissemination of qualitative insights. In recent years, software programs have presented a promising mechanism to accelerate transcription, but the broad application of such programs has been constrained due to expensive licensing or “per-minute” fees, data protection concerns, and limited availability of such programs in many languages. In this article, we outline our process of developing and adapting a free, open-source, speech-to-text algorithm (Whisper by OpenAI) into a usable and accessible tool for qualitative transcription. Our program, which we have dubbed “Vink” for voice to ink, is available under a permissive open-source license (and thus free of cost). We assessed Vink’s reliability in transcribing authentic interview audio data in 14 languages, and identified high accuracy and limited correction times in most languages. A majority (9 out of 12) of reviewers evaluated the software performance positively, and all reviewers whose transcript had a word-error-rate below 20% (n=9) indicated that they were likely or very likely to use the tool in their future research. Our usability assessment indicates that Vink is easy-to-use, and we are continuing further refinements based on reviewer feedback to increase user-friendliness. With Vink, we hope to contribute to facilitating rigorous qualitative research processes globally by reducing time and costs associated with transcription, and expanding the availability of this transcription software into several global languages. With Vink running on the researcher’s computers, data privacy issues arising within many other solutions do not apply.

**Summary box:**

- **What is already known on this topic:** Transcription is a key element to ensure quality and rigor of qualitative data for analysis. Current practices, however, often entail high costs, variable quality, data privacy concerns, stress for human transcribers, or long delays of analysis.
- **What this study adds:** We present the development and assessment of a transcription tool (Vink) for qualitative research drawing upon an open-source automatic speech recognition system developed by OpenAI and trained on multilingual audio data (Whisper). Initial validation in real-life data from 14 languages shows high accuracy in several languages, and an easy-to-use interface.
- **How this study might affect research, practice or policy:** Vink overcomes limitations of transcription by providing a ready to use, open source and free-of-cost tool, with minimal data privacy concerns, as no data is uploaded to the web during transcription.

## Introduction

Recent decades have witnessed an ever-increasing use of qualitative approaches in global health research (1, 2), due at least in part to a recognition that in-depth, qualitative insights can add richness to existing data and can facilitate more person-centered, ground-up solutions to health challenges (3). A factor that limits broader and timelier use of qualitative data is transcription. Transcription refers to the process of converting recorded audio speech, from an interview or focus group discussion, into a written format. Transcription is an indispensable part of the qualitative process. The selection of an adequate transcription approach (e.g. transcribing dialogue versus also capturing utterances such as “uh-huh” or “umm”, details of who is speaking, interruptions, pauses, or involuntary and non-lexical noises such as coughs or throat clearing) is seen as crucial to maintain quality and rigor of data (4, 5), yet the processes and decisions made during transcription represent an often neglected space within qualitative scholarship, receiving limited attention and reporting in the literature. A recent review about reporting of transcription processes found that 41% of articles employing interviews as a research method did not mention transcription, while 11% mentioned transcripts but not the process of transcription (6). Given the extensive use of transcription in qualitative research, the limited discourse on the processes, strengths and limitations inherent to transcription is striking (7).

To date, transcription has mainly been accomplished in three ways: by a single researcher or research team who listens to the audio files and manually types text; by professional transcription services wherein recorded material is sent to a company that then returns transcripts; or by software-based transcription programs that entail payment to an external platform where recorded material is uploaded, automatically transcribed (with or without additional accuracy checks) and transcripts can then be downloaded. Each of these existing approaches entails opportunities and challenges. Manual transcription by the lead researcher or team facilitates extensive engagement with the data, but it is time consuming for the individual(s) transcribing and for the project as a whole. One hour of recorded material typically requires six to seven hours of transcription time (8). Despite being inherent to the process of manual transcription, delays can lead to collected data waning in relevance (9) or, as witnessed in COVID-19 (10), becoming obsolete. Many qualitative teams have sought to mitigate transcription delays by forgoing verbatim transcription in favor of selective transcription or via capturing data in the form of field notes and summaries (11, 12). While selective transcription and related techniques can facilitate timely results, these approaches can increase the risk for researcher bias and information loss (13).

Increasing the number of individuals transcribing a dataset by outsourcing transcription can reduce time but may increase project expenses (14) and cause variability of transcript quality and content, as transcribers may have little familiarity with the research aims (15). Additionally, in case of emotionally straining research topics or respondent narratives, outsourcing can induce mental stress for transcribers who otherwise would not have come in contact with the data (16). Data safety and privacy are also a concern when sharing raw data with individuals outside the study team.

Software-based alternatives (e.g., NVivo, TranscribeMe, happyscribe, OneNote (Microsoft) or Smart Pen (17)) are new entrants into the transcription field whose broad utility in academic research has been limited by a few factors (18). In some cases, programs require training on a user’s voice, which is a time-consuming step that reduces the program’s sensitivity to other voices (19). In other cases, software-based services are expensive and exclusionary, which hinder their use in projects with limited funding or in projects that use languages that transcription firms do not place within their range of products (20). Literature on the consistency and accuracy of speech-to-text software is currently limited, but at least one study showed that accuracy varied widely depending on the used algorithm and decreased overall with audio files that were low-quality or entailed multiple speakers (21). This presents further challenges for researchers since qualitative data often stems from conversational speech (e.g., interviews, focus group discussions wherein multiple speakers and background noise are common). Since software developers often don’t provide word-error-rate for this sort of non-naturalized audio recordings, further exploration in this field is necessary (22).

In response to the existing challenges of cost, timeliness, availability, exclusivity and reliability, and with the advent of stronger and less resource-intensive algorithms for everyday use, software engineers and computer scientists worldwide have begun debating feasibility, trade-offs, and opportunities related to transcription via open-source (i.e., free-of-cost) speech-to-text algorithms. Such a platform would mitigate several barriers inherent to manual and/or commercial transcription, but as of now we are not aware of a program that is adjusted to the needs of qualitative researchers, is user-friendly in terms of navigation and is available in an equitable format in terms of language, downloadability and cost.

In this article, we outline our process of developing and adapting a free, open-source, speech-to-text algorithm into a usable and accessible tool for qualitative transcription. We assess our standalone application ‘Vink’ for reliability in transcribing authentic interview audio data in several languages, and we provide a detailed step-by-step guide for researchers considering using this tool for their own data transcription.

## Developing and testing a free transcription package

### Development

As a first step in developing our transcription tool, we identified open-source speech-to-text (STT) algorithms including VOSK by Alpha Cephei (23), Silero (24) and Whisper by Open AI (25). These algorithms were pilot tested using real-life interview data in German in an exploratory approach. Whisper by OpenAI (25) (see breakout box 1) was selected as the best option based on the accuracy and readability of transcripts, the inclusion of punctuation as well as upper and lower case lettering in the resulting transcript, robustness to background noise and the program’s potential applicability in numerous languages.

#### Breakout Box 1

**Whisper by OpenAI**

Whisper by OpenAI is an open-source automatic speech recognition system (ASR) trained on multilingual audio data in an end-to-end approach. OpenAI emphasizes its ability to navigate transcription that captures or mitigates challenges related to accents, background noise and technical language. The algorithm uses one single speech model that automatically recognizes the audio file language and transcribes the data. Audio recordings with mixed languages can therefore also be transcribed easily. Since Whisper was not built via one specific dataset or voice, the system is applicable across qualitative, global health research projects. Furthermore, Whisper runs locally on the user’s computer without requiring a data upload, thereby mitigating privacy concerns. While the program does not require an online connection, running Whisper requires good hardware as it uses between 1-10 GB of RAM, depending on which of the five available speech model sizes is selected. Using Whisper thus entails a trade-off: if a higher level of transcription is sought, the program’s runtime and necessary RAM will increase.

Like many currently available ASR algorithms, Whisper requires knowledge of software programming (e.g. Python), placing it beyond reach for researchers who lack programming skills. Noticing this gap, we developed a standalone application to make the potential utility of Whisper available to a broader pool of researchers. Our goal was to create a downloadable, ready-to-use transcription package that bundles the Python interpreter, the Whisper Python package as well as all its dependency into one tool that allows you to run the Whisper algorithm on one’s personal computer. We also wanted a product that had an easily navigable user interface and could be free to anyone interested in using it for their own research.

The final transcription tool, which we dubbed “Vink” due to its ability of transferring textual data from voice to ink, is available at https://heibox.uni-heidelberg.de/f/6b709d18b0d244cdb792/ (FOR REVIEW PURPOSES, FINAL WEBSITE TO BE INSERTED UPON PUBLICATION). More technical information on this standalone application, which was created using PyInstaller, are available online at https://github.com/ssciwr/whisper-standalone/. The tool currently is only available for Windows; development of macOS and Linux versions is in progress. All assessment was done anonymously and did not include any personal or individually identifiable information. The institutional review board of the medical faculty, University of Heidelberg, Germany, therefore exempted this study from ethical review.

### Assessing the reliability of Whisper on multilingual realistic audio data

We assessed the performance of the Whisper algorithm when transcribing realistic audio data in 14 languages including: American English, Arabic (Classical Arabic), Bahasa Indonesia, Burmese, Chinese (Mandarin), Filipino, French, German, Malagasy, Portuguese (Brazilian), Spanish (Colombian), Tamil, Turkish, and Yoruba. Multilingual transcription reviewers provided audio data of a discussion in their mother tongue following detailed recording instructions (see Appendix 1). To mimic real-life qualitative data quality, audio files were recorded on either a phone or a regular recording device in a quiet setting. Audio quality was controlled by the lead author. Transcripts of the audio files were generated using the medium size language model of Whisper (5GB RAM required) and were sent back to the reviewers for assessment. Reviewers were then asked to correct the automatically generated transcript in one sitting, and to record the time needed to correct the transcript and the word error rate (WER) including errors linked to the deletion of filler words (e.g. “uhh” or “umm”). For reviewer instructions see Appendix 2. Reviewers were then asked to complete an anonymous questionnaire on the perceived usefulness of the transcript (see Appendix 3). Study data were collected and managed using REDCap electronic data capture tools hosted at the Universitätsklinikum Heidelberg (26, 27).

Following this approach, a total of 19 audio files were provided, 14 of which were assessed. The remaining 5 reviewers did not provide an assessment of the transcript (3 contact reminders were made).

### Reliability and perceived usefulness of the generated transcripts

Table 1 summarizes the recordings assessed and the algorithm’s transcription performance. Substitutions describe replaced words (e.g. transcribing “house” for “mouse”). Deletions were cases in which words or non-verbal cues were left out of the transcript, and insertions represent added words that were not said.

**Table 1.**
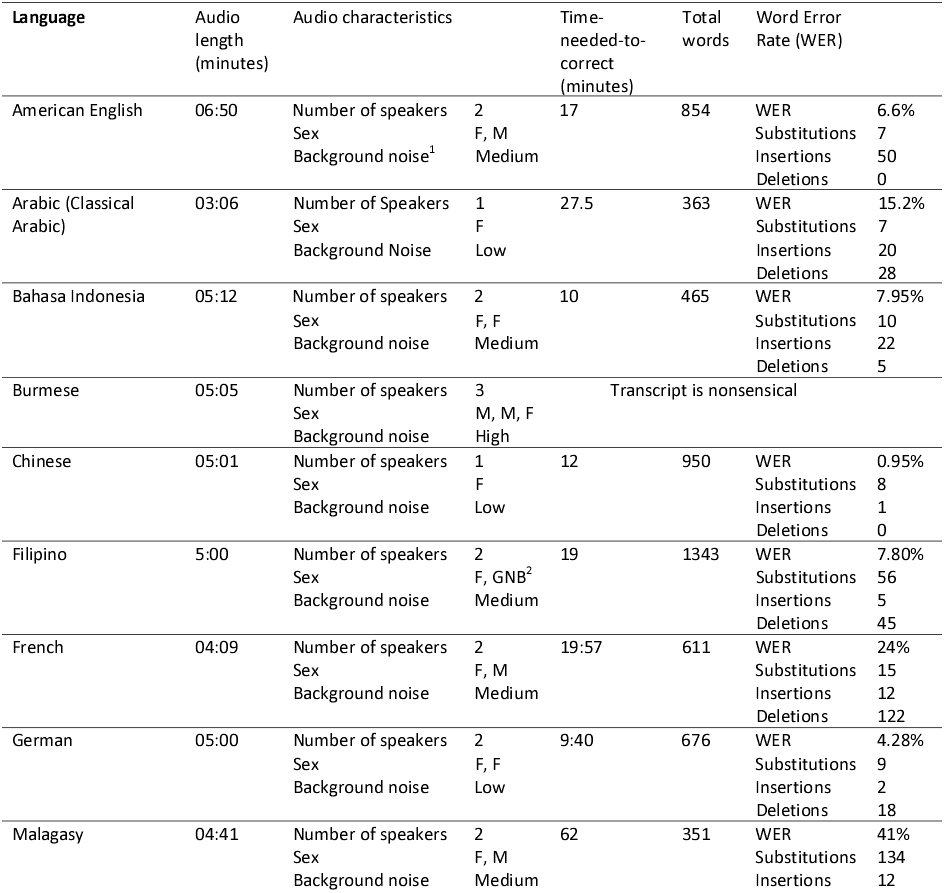
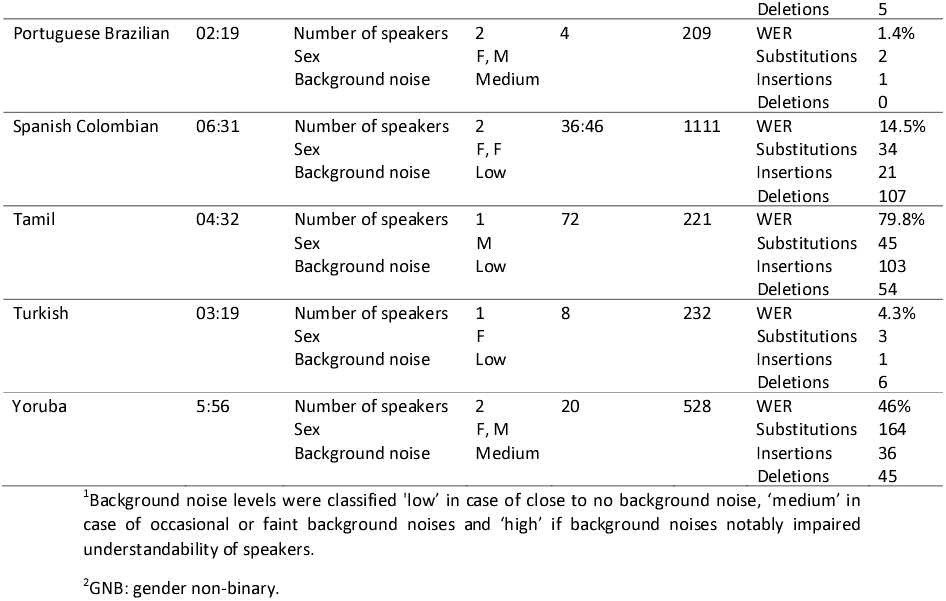
Word Error Rate and time-needed-to-correct of Vink-generated transcripts of audio data

As expected, the performance of Whisper varied widely across languages, with Tamil and Burmese producing the least useful transcripts. The tool’s performance did not seem to be language group specific with e.g., high accuracy in Chinese (Mandarin) and extremely low accuracy in Burmese. More likely, this is correlated to the very low percentage of e.g. Burmese audio in the training dataset of the Whisper algorithm ((25) Appendix E). Among European languages, French required the most extensive transcription correction.

The time needed to correct transcripts varied greatly. Controlling for the different lengths of audio recordings, it resulted in 1.7-fold (Portuguese) to 16-fold (Tamil) the length of the original audio.

Overall, the generated transcripts were evaluated positively. All reviewers whose transcript had a WER below 20% (n=9) indicated that they were either likely or very likely to use Vink in their future research. However, the results from the questionnaire revealed several areas for improvement.

The algorithm seems to naturalize the text output and therefore rarely includes filler words in the transcript. Non-verbal vocalizations such as laughing, crying or hesitations are omitted as well. Repetitions are partly cleared in the final transcript, producing a denaturalized transcript version (28). The deleted non-verbal vocalizations account for a significant part of the WER in our assessment. For instance, the algorithm would naturalize the sentence “We, ehm, wanted to gi-… give an example.” To “We wanted to give an example.”, which would be counted as two deletions in our assessment. Respondents wished for hesitance and pauses to be included and captured with an ellipsis symbol (“…”) rather than a comma.

According to respondents, the algorithm (as described in previous papers on ASR (21, 29)) struggled during crosstalk segments of the audio data. Some respondents suggested that underlining the pause between sentences in the audio recording could be helpful, for instance line breaks between speakers to make it clear who was speaking. Speaker recognition would be helpful to distinguish the different voices which often is a challenge during focus group discussion sessions.

The perceived readability of transcripts correlated with the WER of the respective transcript, with an overall high readability across languages. In total, 10 out of 12 reviewers indicated that they were either likely or very likely to use Whisper in future research projects.

## Results of the short questionnaire are presented in Figure 1.Usability of Vink

### Testing the usability of our transcription package with user interface

To gauge the usability of Vink, we gave 5 people without previous experience in computational science access to the transcription package and provided them with an instruction sheet (see Appendix 4) on how to download and use the transcription tool. We then observed how well users were able to navigate our transcription tool using cognitive think-out-loud interviewing during first use. In addition, we asked users for feedback regarding how they perceived the tool in terms of usability and user-friendliness, and what changes they suggested to increase usability.

**Figure 1.**
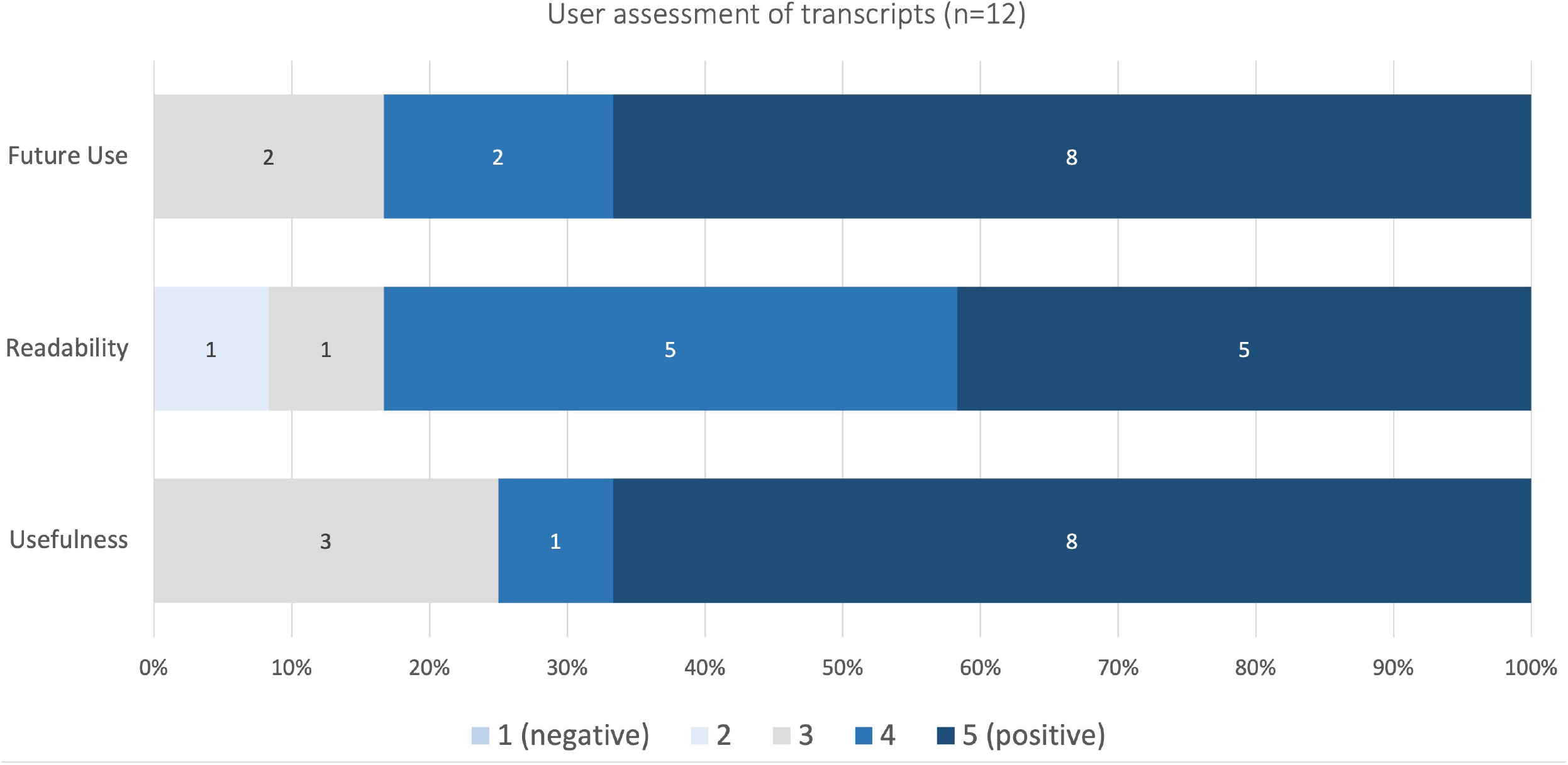
Perceived usefulness, readability of transcripts and likeliness of future use

### Challenges and Improvement

Our usability assessment showed that reviewers were able to install Vink and transcribe an audio file using the included interface. Reported issues included difficulties finding the executable file for Vink in the downloaded folder and confusion about suitable text file formats, which were addressed in the current version of Vink to enhance user friendliness. Inter alia an installer was added to facilitate the set-up process. Most struggles and uncertainties resulted from reviewers overlooking content in the instruction manual, highlighting the importance for our team to maximize the self-explanatory nature of the interface. See Appendix 5 for the complete list reported usability issues and subsequent improvements. Vink’s interface and the instructions for use were also further modified following a rapid, iterative approach that draws upon human-centered design. The user manual to the newest version of Vink can be found in Appendix 6.

### Summative evaluation of Vink

Taken as a whole, existing standards for transcription present challenges that can be addressed via ASR algorithms and standalone applications such as Vink. Table 2 summarizes overarching challenges to traditional verbatim transcription, outlines how Whisper as an ASR algorithm can address some of these challenges, and details the characteristics and resulting additional needs of our transcription tool Vink.

**Table 2.** Needs of traditional transcription, opportunities via Whisper and opportunities via Vink.

| Transcription concerns and needs | Characteristics of Whisper | Characteristics of Vink and additional needs |
| --- | --- | --- |
| **Resources, infrastructure, and costs** |  |  |
| Manual transcription is a time-consuming process. | Automatic transcription potentially reduces the time to generate text from audio files. | Review and correction of transcripts is required (a step also recommended for manually generated transcripts) |
| Transcription services are expensive. | Whisper is offered by OpenAI free of cost. | Vink is a free of cost transcription tool using Whisper's open-source algorithm. |
| Transcription software often requires high computing power to operate. | Whisper offers multiple model sizes that require 1-10 GB of RAM, thus can run on average computers, depending on the model size. | Vink conserves this feature from Whisper, allowing selection of model size per user and computer characteristics. |
| **Safety and privacy** |  |  |
| Uploading data for transcription or outsourcing transcripts to a third party raises confidentiality and data protection issues. | Whisper runs locally, thus eliminates the need to share or upload data. | Vink is designed to operate locally without uploading data. |
| **Quality of transcription** |  |  |
| Transcription software is often unavailable in non-Western or less dominant languages. | One model for all languages technically makes Whisper usable for everyone. Audio files with mixed languages can be transcribed. | Accuracy of transcription varies across languages (Table 1). |
| Conventional transcription software often requires training on a user's voice or on exemplary audio data. | The Whisper algorithm has already been trained on big data and is ready for use. | The 'ready to use' feature limits the possibilities to adapt the algorithm to individual requirements. |
| Conventional transcription software often struggles with accents, mixed use of languages and background noise. | Whisper provides improved robustness to accents, background noise and technical language. | The improved speech recognition comes at the expense of expressions (e.g., laughter) that are excluded from the final transcript. |
| Identifying speakers (e.g., interviewer, respondent, multiple participants) is an essential but sometimes challenging feature of transcription. | Whisper does not offer speaker recognition. | Vink currently does not include speaker recognition. Depending on the transcription approach, the user may need to add them manually. |
| Other open-source transcription software (Silero, Vosk) only output raw lower-case text. Punctuation models can be applied later in the process, but these are not available for all languages. | Whisper generates transcripts with already integrated punctuation and upper cases regardless of the language. |  |
| **Ease of use** |  |  |
| Transcription software should be accessible to researchers without knowledge of software programming. | Whispers requires a programming language (e.g., Python, R), an interpreter and installation of specific packages within the programming software, to operate. | Vink is a downloadable standalone application which includes the necessary packages and tokenizers, reducing the installation requirements and steps. |
|  | Whisper does not have a user interface, which limits its use to people with knowledge of programming (e.g., Python). | Our transcription tool includes an intuitive user interface. |

## Discussion

Vink is an easy-to-use, open-source speech-to-text tool for qualitative research that is free of cost, making it an accessible transcription solution for research projects. Vink’s accuracy in non-western and (in a research sense) rarer languages, as well as the limited computing power required to operate it, make our transcription tool usable for everyone with access to a standard computer or laptop. These characteristics may help mitigate global disparities in health research resources (30). In addition, compared to uploading data to third-party transcription services, Vink runs locally, which allows protection of privacy and confidentiality of data, an established principle of qualitative research (31, 32).

The accuracy of generated transcripts is central to the application’s value in qualitative research. Poland (33) defined transcription as accurate according to its faithfulness to the original speaker’s intention and its fit with the research aims. In practice, transcripts are often considered accurate when they match the recorded audio, disregarding the original interaction. Although problematic as this takes a purely positivist view that there is one ‘correct’ version, this understanding allows for a comparison of transcripts and presents a feasible common ground for accuracy assessment in our case. Part of this consideration on transcript accuracy is the inclusion of behavioral annotations. Gestures and non-verbal vocalizations can be considered representative of e.g., the speakers’ engagement in the interview or topic or certainty with expressed opinions. However, non-verbal cues are often excluded from transcripts, whether those transcribed by hand or via the support of an algorithm. This form of ‘selective transcription’ of the data increases readability but loses data and risks researcher bias. By virtue of saving time on the documentation of words, Vink may allow more time to capture and annotate the broader context of the interview or focus group engagement.

In our descriptive and Radford’s (25) large scale assessment of Whisper’s accuracy on multilingual speech, the overall performance (or word-error-rate (WER)) of the algorithm is good. Variability in WERs show that despite the algorithm technically being applicable to a high number of languages, disparities in accuracy remain across languages, commonly favoring languages such as English, German, and Chinese. In a few languages that are linguistically further from English or for which the amount of audio data used in training Whisper was comparatively low ((25) Appendix E), the quality and therefore usefulness of the transcripts decreased. While the amount of respective audio data for training is strongly correlated with Whisper’s performance, an additional factor for those languages is a lack of transfer due to the linguistic distance from the predominantly English audio data (65%).

The lack of transparency regarding metrics in machine learning literature (34), including the exact definition of the WER in the original publication on the Whisper algorithm (25) challenge comparisons across programs. For example, it is not clear whether filler words are considered in the WER assessment. Such deletions are relevant for qualitative research, as most errors were due to deletions of non-verbal vocalizations, which might be of importance since pauses can indicate divided attention or nervousness of the interviewee (35). The WER as a metric does not account for the causes of errors. Factors that can affect WER, independent of the capabilities of the ASR technology, include recording quality, technical terms or proper nouns, background noise, sex of the speaker, pronunciation and speech fluency. These might explain the differences in WER between our own assessment and the large scale original assessment of Whisper’s WER (25). With the limitations of the WER, other means (e.g., perceived usefulness or time-needed-to correct) provide valuable information for a realistic assessment of the transcript’s value for researchers. In our findings, the readability of transcripts was perceived as high, which implies an accelerated process of correction since the text can be followed and adjusted more easily.

Researchers have argued that computers may tempt qualitative scholars to perform ‘quick and dirty’ research (36) and could lead to a loss of closeness to the data (37). In the context of automated transcription, we see the risk of generated text being superficially evaluated in terms of its readability and not by its nuanced representation of the original recording, including non-verbal cues. Additionally the Whisper algorithm is trained to condition on the history of text of the transcript in order to use longer-range context to resolve ambiguous audio (25). Sentences with non-understandable parts are reconstructed leading to overall higher accuracy and good readability but possibly a false sense of certainty of transcript correctness in hard-to-understand passages. We therefore advocate for researchers considering using speech-to-text algorithms (including Vink) to carefully choose its exact mode of application. Especially for researchers interested in nuances of human interaction, too much reliance on the automatically generated transcript might cause a significant loss of valuable data. The applicability of automated transcription is also challenged by scholars such as Lapadat (38) who views transcription as a process rather than a product, as it involves constant decisions regarding how to present the data and which additional information to include. This makes transcription an inherently interpretative act, influenced by the transcriber’s own biases and assumptions (39). As algorithms are not able to make such decisions about meaning-making and interpretations, nor about ways in which these meanings may best be represented (21), we propose that ASR generated transcripts should merely be seen as a first step in the transcription process, which are to be revisited and modified (40).

In terms of limitations, Vink is currently only available for Windows computers, which restricts its potential user base. We are working on a macOS and a Linux version. Another limitation of our study is the varying levels of experience in transcribing or correcting qualitative data from our reviewers. We did not account for the experience of reviewers or the use of tools for correction, which may have introduced variation in the time-needed-to-correct and WER assessments. Larger, systematic evaluations of the software may overcome this limitation. Similarly, we did not assess the algorithm in several contexts relevant for qualitative research (e.g., focus group discussions, speech with strong accents, more background noises). As qualitative research often is performed in settings where the researcher only has limited control over environmental factors, such further assessment would allow a firmer establishment of the conditions required for the software performance to be sufficiently useful.

### Going forward

A step-by-step guide on how to install and use Vink is available for use (Appendix 6). The code for the graphical user interface of Vink, as well as the combined work with the bundled dependencies are published under the MIT license. Vink’s installer will also install a number of bundled software packages under a variety of software licenses (Nvidia License Agreement for Nvidia SDKs, LLGPL v3, MPL v2, PSF License, Apache 2.0, BSD-3, BSD-2, MIT, Zlib license, Unlicense). For detailed information about these licenses, please read the license agreement. We ask users to credit OpenAI when using the algorithm, and to cite this publication when using Vink in their own work. As mentioned, Vink-generated transcripts should be seen as a first step in the transcription process, which are to be revised by research teams (and ideally, those who undertook the data collection activity and/or who will undertake data analysis).

We are happy to hear about other researchers’ experiences, successes, and challenges in applying this approach to automatic transcription in their own work and are open to feedback and suggestions. A portal for feedback will be available on the website where Vink can be downloaded. Additional guidance and information on the Whisper algorithm are available online (not moderated by us), for example at https://openai.com/research/whisper or https://github.com/openai/whisper. Tutorials and forums to chat about possibilities and limitations of automated speech-to-text transcription are emerging, allowing for an exchange between interested individuals. To the best of our knowledge, such forums are primarily technical in nature.

We aim to improve and update the standalone package in the future. Improvements in language models will be considered in newer versions. Being an open-source algorithm means that the way this program operates is more transparent than in commercial software and can be examined by the research community.

## Conclusion

In this article, we have introduced and evaluated our novel transcription tool Vink for automated interview transcription in various languages, based on OpenAI’s Whisper. Our findings outline the possibilities of integrating open-source speech-to-text algorithms into qualitative research. With the current rapid developments in this field, we expect the accuracy, relevance and ease of use of ASR to continue to increase and want to contribute to the emerging discourse on its resulting potentials and drawbacks for qualitative research. We hope that by providing a ready-to-use and free tool, we will allow qualitative researchers, especially those with limited resources, to save time and money. This in turn might be reinvested in engaging more profoundly with data and deepening other steps of the analytic process, thereby ultimately strengthening the quality of qualitative research across settings and disciplines.

## Competing Interests

The authors declare no competing interests.

## Data Availability

Additional data are available upon reasonable request and after approval of our IRB
Out of privacy concerns we did not share the audio recordings, nor the original and corrected transcripts of these audio recordings. This also applies to the audio recordings of the usability assessment.
To request access please contact the corresponding author Hannah Tolle.

## Appendices

1. Information sheet transcription reviewers
2. Instructions for assessment of generated transcripts
3. Short questionnaire perceived usefulness
4. Instructions for use of the transcription tool
5. Results usability assessment
6. Updated User Manual for Vink

## Acknowledgments

We thank to all the reviewers that contributed to the assessment of the Vink app’s usability and evaluation of transcripts. They include Lukas Brümmer, Myo Chit, Abeer Fandy, Zavaniarivo Rampanjato, Mark Donald C. Reñosa, Sonjelle Shilton, Girish Srinivas, Anete Trajman, Stefan Weber, Rayan Younis, among others that preferred to remain anonymous.

We would like to thank the Scientific Software Center of the Heidelberg University for their development work on this project. The Scientific Software Center is funded as part of the Excellence Strategy of the German Federal and State Governments.

We thank Frank Tobian for the technical support of this work.

We thank the team from FIND for their support.

## Notes

### Competing Interest Statement

The authors have declared no competing interest.

### Funding Statement

Shannon McMahon, The Scientific Software Center Heidelberg (funded as part of the Excellence Strategy of the German Federal and State Governments), the
National Institute of Allergy and Infectious Diseases, NIH, USA (U01AI152087) for the Rapid Research in Diagnostics Development for TB Network (R2D2 TB Network) as well as the Ministry of Science, Research and the Arts Baden-Wuerttemberg

### Author Declarations

The reviewers of the automatically generated transcripts did only submit technical information about the transcript. They did not provide any personal information. All assessment for perceived usefulness was done anonymously and did not include any personal or individually identifiable information. The assessment for usability of the application was done within the research group. The institutional review board of the medical faculty, University of Heidelberg, Germany, therefore exempted this study from ethical review.

