## Appendix 1 for "From voice to ink (VINK): Development and assessment of an automated, free-of-charge transcription tool"

### Transcription Software Evaluation

Thank you again for signaling potential interest in contributing audio recordings to support the evaluation of a novel, free transcription approach. As discussed, we would like to give you some more information on what this evaluation is about and what your participation would entail.

#### What is the aim of this project?

We would like to assess the performance of a new, open-source transcription software (whisper) in transcribing authentic audio data in different languages. The successful use of free transcription software could make a big difference towards making qualitative analysis less resource and time intensive, especially in a global health perspective.

#### Which languages are we interested in?

In our first evaluation we would like to focus on some rather common languages that are of interest in global health research. Therefore, we are searching for volunteers who speak ***English, French, Spanish, Portuguese, Swahili, Filipino, Arabic or Chinese***. However, we are also interested in expanding this list, depending on the availability of audio recordings– please reach out to Agrin (see below) if you would like to offer another language.

#### What would your contribution entail?

We would kindly ask you to ***record a discussion with a friend or family member in your native language***. This audio recording should ideally be about 3-5 minutes long. If it is possible, we would prefer having recordings with a male and female voice.

To ensure realistic quality of the audio data we would prefer it if you could record your discussion on either a phone or a regular recording device in a rather quiet setting without distractions or too much background noise. Please don't eat or drink while making the recording. However, if you absolutely cannot avoid having some noise on your recording this is fine as well.

We would ask you to talk as freely as possible. Please don't try to speak extra clearly, since we are particularly interested in authentic real-life audio data. ***We want to highlight that we will not analyze any content of the shared audio files, nor will the transcript be shared with anyone besides the person running the software and the person evaluating it (ideally you, see below) without your explicit prior approval. Transcripts and audio files will be destroyed immediately after evaluation is complete***. Please do not be overly conscious of what exactly you discuss – it can be as mundane as you want, as long as it has a similar flow to a normal conversation.

**Ideas for discussion topics:**

However, to make the discussions easier for you and to ensure a comparability of the language level of the different recordings, we would suggest you talk about one or several of the following topics:

- What does global health mean to you?
- How does your work relate to global health?
- Which experiences have you made with qualitative data collection?
- Which global health fields are you most interested in and why?

Once you have finished your recording, ***please submit it to Agrin.***

**What comes after recording and how does the evaluation work?**

Once we have received your recording, we will run the transcriptions software on your audio data. To ensure maximum accuracy and privacy, it would be great if you would also be available to assess the transcription's accuracy of your own recording (we expect this to not take more than 15 minutes of your time). If you are available for this, we would then send the resulting transcript of your recording back to you (most likely within a week of your submission).

We would then ask you to ***please assess how well the automatic transcription performed on your audio data***. We would like you to do this by correcting the transcript (by relistening your recording) and determining the word error rate of the transcription (how many words did the program get wrong in relation to how many words there are in total), as well as an estimation of how long it took you to correct the transcript. However, we would share a more detailed description on the evaluation criteria when sending you the transcript. To conclude we would then ask you to answer 3-4 short questions on how useful you perceived the transcription.

We would be very thankful if you decided to support this project. If you have any questions, please don't hesitate to reach out to Agrin (for organizational questions,) or Hannah (for software/technical questions,).
