## Appendix 2 for "From voice to ink (VINK): Development and assessment of an automated, free-of-charge transcription tool"

Dear Participant,

Thank you very much for having contributed an audio file to our project.

You will find attached the transcript of your audio file which was generated automatically using our transcription application based on Open AI's "Whisper".

As mentioned before, we would like you to assess the transcription's accuracy of your own recording to ensure maximum accuracy and privacy (we expect this to not take more than 15 minutes of your time). Therefore, ***we would like to ask you to correct the transcript that was sent to you.***

Please see below some specific instructions regarding how to assess the transcript accuracy. If you have any questions, please reach out to me.

- Please correct the transcript while relistening to your original audio recording and **measure the time you needed to correct the transcript**. To ensure that the time measured reflects the actual time needed to correct, please correct the transcript in one single sitting. Please feel free to use the transcription approach that you feel most comfortable with and that you would use when transcribing by hand yourself.
- **Word error rate** (this will allow us to make comparisons with other software, open and paid). Please count how many words the transcription software "got wrong", how many it added and how many are missing. For detailed description of how to calculate the WER, please see the explanation below. Please also consider filler words.

**Please enter the results of your assessment (WER and time needed to correct) in the table on top of the transcript and send the corrected version to Agrin.**

To conclude we would then ask you to answer four short questions on how useful you perceived the automatically generated transcript. For this, we have created a fully anonymous questionnaire on RedCap which you can access via the following link:

https://cru.med.uni-heidelberg.de/redcap/surveys/?s=4JAN44XHKDPAJKRL

**Thank you very much for your time and effort!**

**How to calculate the word error rate (WER)?**

The WER is the number of errors divided by the total words, according to the formula:

**Word Error Rate = (Substitutions + Insertions + Deletions) / Number of Words Spoken**

To calculate the WER, please start by adding up the substitutions, insertions, and deletions that occur in a sequence of words in the transcript, as described below:

- A **substitution** occurs when a word gets replaced (for example, “noose” is transcribed as “moose”)
- An **insertion** is when a word is added that wasn’t said (for example, “SAT” becomes “essay tea”)
- A **deletion** happens when a word is left out of the transcript completely (for example, “turn it around” becomes “turn around”)

Then, divide that number by the total number of words originally spoken. The result is the WER.

This can be illustrated with an example[1]:

Let’s say that a person speaks 29 total words in an original transcription file. Among those words spoken, the transcription included 11 substitutions, insertions, and deletions.

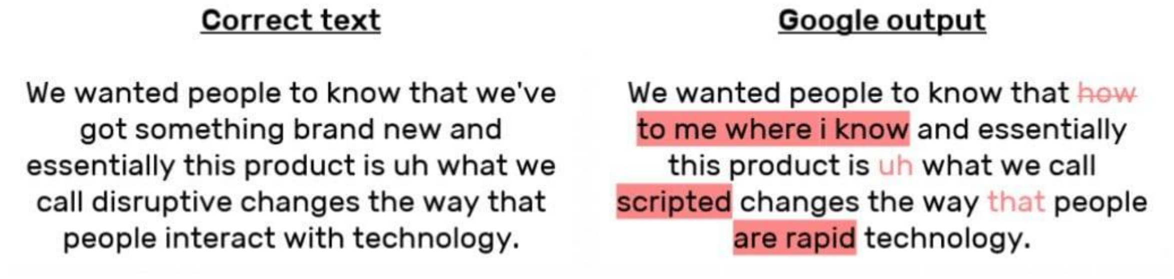


To get the WER for that transcription, you would divide 11 by 29 to get 0.379. That rounds up to .38, making the WER 38 percent.

[1] Taken from: https://www.rev.com/blog/resources/what-is-wer-what-does-word-error-rate-mean#:~:text=Basically%2C%20WER%20is%20the%20number,The%20result%20is%20the%20WER.
