## Appendix 3 for "From voice to ink (VINK): Development and assessment of an automated, free-of-charge transcription tool"

### Whisper Transcription Project

Dear Participant,

As a last step we would like to ask you to please complete the questions below.

Thank you for your effort!

---

1) What was the Word Error Rate (in %) of your transcript?

- ◯ 0-10
- ◯ 11-20
- ◯ 21-30
- ◯ 31 or more

---

2) How useful did you find the use of the automatically generated transcript compared to manual transcription?

- ◯ very unuseful
- ◯ unuseful
- ◯ undecided
- ◯ useful
- ◯ very useful

---

3) Please judge how ‘understandable’ the transcript is.
See an example below:

Spoken: "I like to bike around"
Model 1 prediction: "I liked to bike around"
Model 2 prediction: "I like to bike pound"

In the above example, both Model 1 text and Model 2 text have a WER of 20%. But Model 1 clearly results in a more understandable transcription compared to Model 2. That’s because even with the error that Model 1 makes, it still results in a more legible and easier to understand transcription.

- ◯ very incomprehensible
- ◯ incomprehensible
- ◯ undecided
- ◯ understandable
- ◯ very understandable

---

4) How likely are you to use this transcription software in your future research?

- ◯ very unlikely
- ◯ unlikely
- ◯ undecided
- ◯ likely
- ◯ very likely

---

5) In your opinion, what would need to be improved in the automatically generated transcript?

________________________________________
