## Appendix 4 for "From voice to ink (VINK): Development and assessment of an automated, free-of-charge transcription tool"

### Instructions for installation and use of the transcription application

#### 1. Installing the application

In order to install the needed transcription software on your computer please visit the following website: https://github.com/ssciwr/whisper-standalone#readme

The solution is currently available for Windows 10+, a MacOS solution is under development.

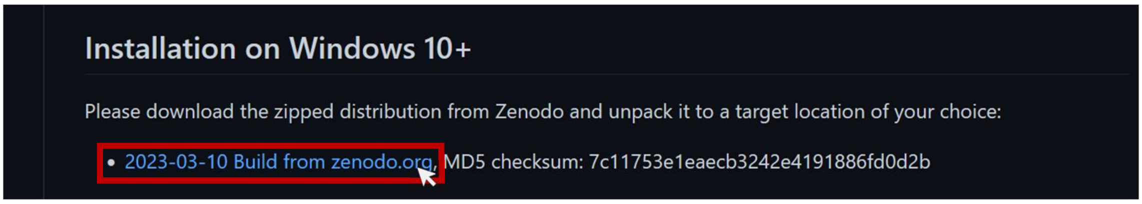


Please download the zipped application folder. Depending on your internet download speed, the download might take a while. Once downloaded, unpack the folder to your chosen target location on your computer.

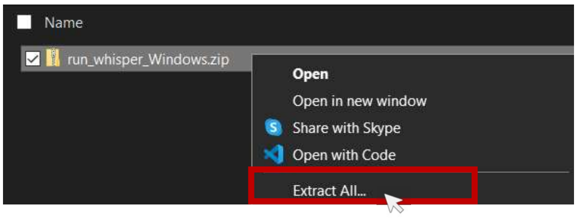


#### 2. Use of the application

Open the unpacked application folder. Scroll down until you see the file "**run_whisper.exe**".

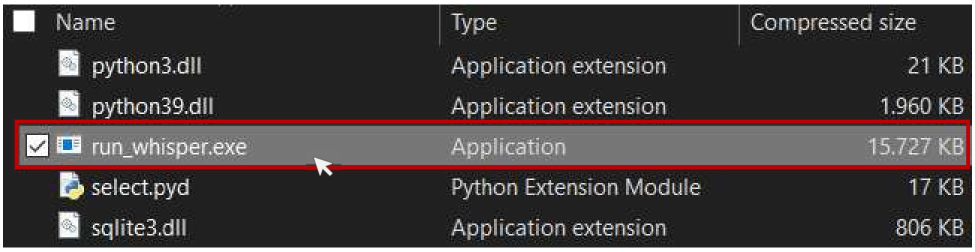


Double click on it and wait until the following user interface opens in the middle of your screen. It is now ready to use.

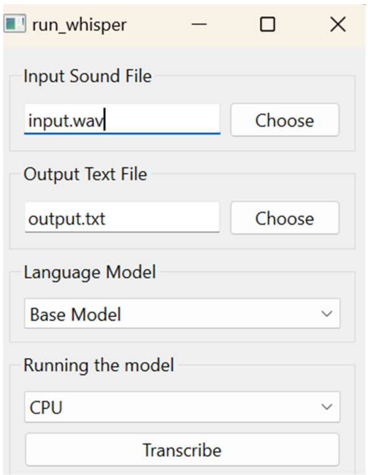

To transcribe the audio file of your choice, please

1. ***Choose the audio file*** (in mp3, mp4, acc, mpeg, 3gpp or wav format) by clicking on the corresponding “choose” button.
2. ***Choose the text file*** (in .txt format) into which you want the application to save the generated transcript. The text file does not generate itself; it has to be created before use.
3. ***Choose the language model size*** that you would like to use for transcription. All models (tiny, small, base, medium, large) run on all languages. With increased model size, accuracy, runtime and required RAM increase as well. If the bigger models are not available, this is because your computer does not have sufficient RAM. You might have to close other programs that are running in parallel.
   Only the smaller models up to base model are included in the first download. If you choose a bigger model for the first time, run time will be somewhat longer because the model has to load before the program starts transcribing.
   There is a special version for transcription in English.
4. ***Choose whether to run the program on your CPU or graphic card***. This will only be necessary if you have a graphic card in your computer. If you have the choice always choose the graphic card for faster transcription.
5. ***Press “Transcribe”***

A progress bar will appear while the program is transcribing the audio file to text. Once the transcription is finished you can find the generated transcript in the chosen text file.
