## Appendix 5 for "From voice to ink (VINK): Development and assessment of an automated, free-of-charge transcription tool"

Supplementary table. Results of usability assessment.

| **Reported usability issues** | **Type of change** | **Description of changes made** |
|---|---|---|
| Not enough memory space to download Vink;<br>Slow download process | Instructions | Required memory space and download time was clarified in the installation process |
| Not enough RAM for larger models due to too many programs running in parallel | Instructions | Included advice to close other programs that are running in parallel on the PC/Laptop |
| Search for the .exe file in the downloaded folder to open the application | Modification of the tool | We added an installer to the application |
| Computer warning about first execution of application | Modification of the tool | Purchase of a code signing certificate; no more need for a firewall exception |
| A pop up of Windows system window appears before the interface | Instructions | Explanation added to instructions |
| Confusion about accepted audio file formats | Instructions | The need of an audio file and required formats are described more prominently |
| Confusion about required text output file and format | Modification of the tool | Application outputs generated text into the interface |
| Confusion about differences between language models | Instructions | Trade-offs between models are explained in more detail |
| Confusion about choice between CPU and Graphic Card | Instructions | Benefits of using each option are now explained |
| Confusion about whether the app works offline | Instructions | It is highlighted that the app needs an internet connection to load models |
