## Appendix 6 for "From voice to ink (VINK): Development and assessment of an automated, free-of-charge transcription tool"

### Manual for installation and use of Vink

#### 1. Installing Vink

In order to install the needed transcription software on your computer please visit the following website: https://heibox.uni-heidelberg.de/f/afbab8dc8d564c69b9de/

The solution is currently available for Windows 10+, a MacOS and a Linux solution are under development.

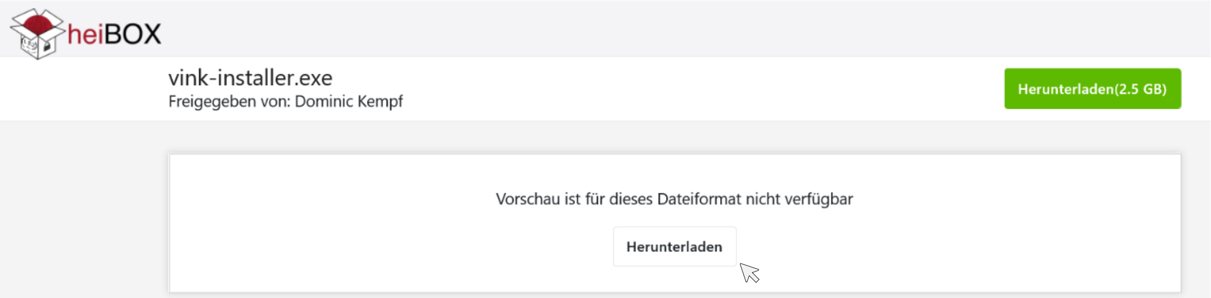


**Please click "Herunterladen" (Download).**
*You will need 4.4GB of storage space to install and run Vink. Depending on your internet download speed, the download might take up to 15 minutes.*

**Once downloaded, double click on the .exe file to start the installation process**.
*It is possible, that your computer warns you about the execution of the unknown file. Please allow for Vink.exe to be run on your computer anyway.*

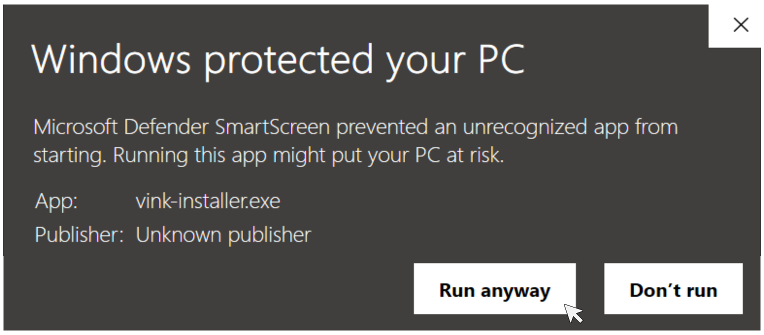


**The installer will appear as a pop up with the License Agreement.** Please read the license agreement and click 'I Agree' to continue the set up.

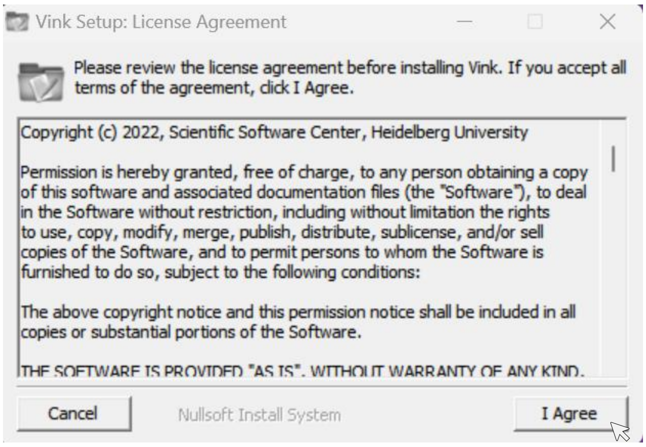

Next, you can choose where to install the application. Per default, the installation will select your computer's Program Files folder.

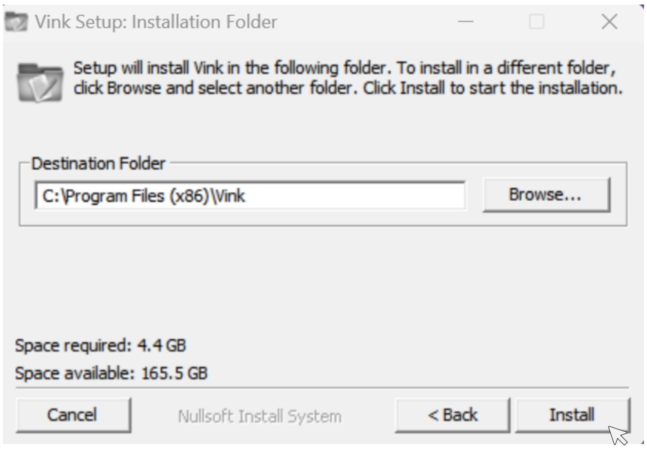


Wait until the Set Up is completed. Vink is now ready to use.

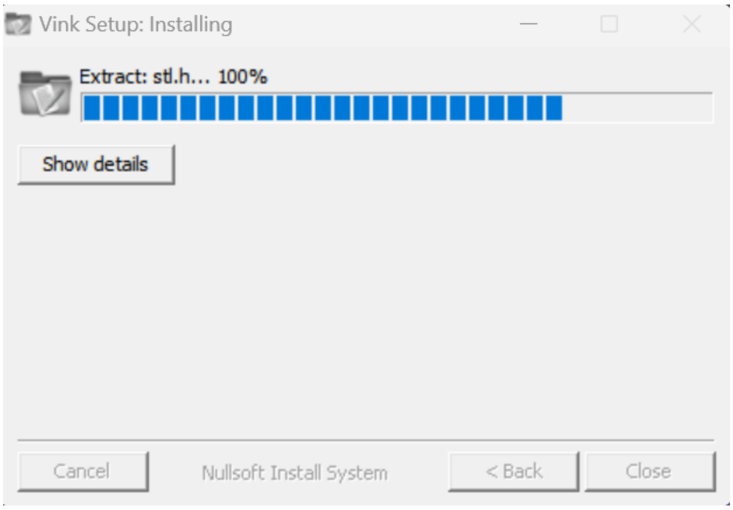

#### 2. Use of the application

Open Vink with a double click.

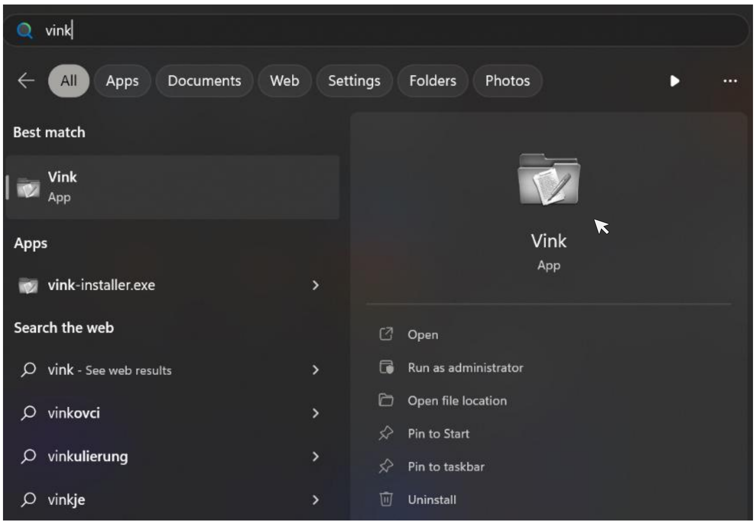


Wait until the following user interface opens in the middle of your screen. It is now ready to use. *Before the interface appears, the black Windows Console will open on your computer. Please ignore this window and wait for the interface.*

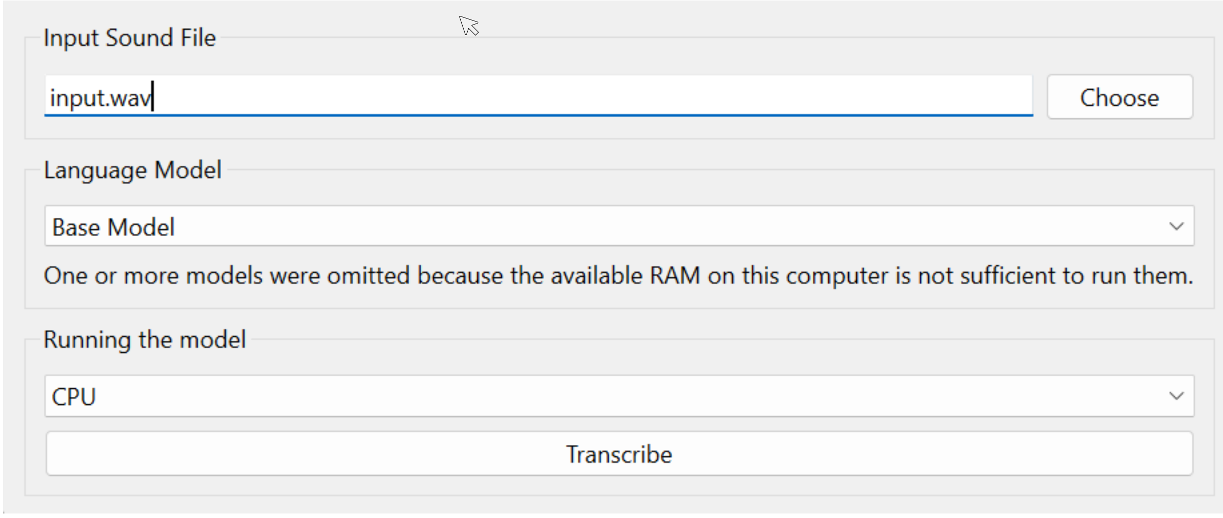


**To transcribe the audio file of your choice, please make sure that you have an internet connection.**

1. **Choose an audio file** that you want to transcribe by clicking on the “choose” button. Accepted formats: mp3, mp4, acc, mpeg, 3gpp or wav format.
2. **Choose the language model size** that you would like to use for transcription. All models (tiny, small, base, medium, large) run on all languages. With increased model size, accuracy, runtime and required RAM increase as well.

   *If the bigger models are not available, this is because your computer does not have sufficient RAM to run them. You might have to close other programs that are running in parallel.*
   *There is a special version for transcription in English.*

3. **Choose how to run the program.** If you don’t have a suited graphic card in your computer, the program will default to CPU. If you have the choice always choose the graphic card for faster transcription.

4. **Press "Transcribe"**

A progress bar will appear while the program is loading the model and transcribing the audio file to text.

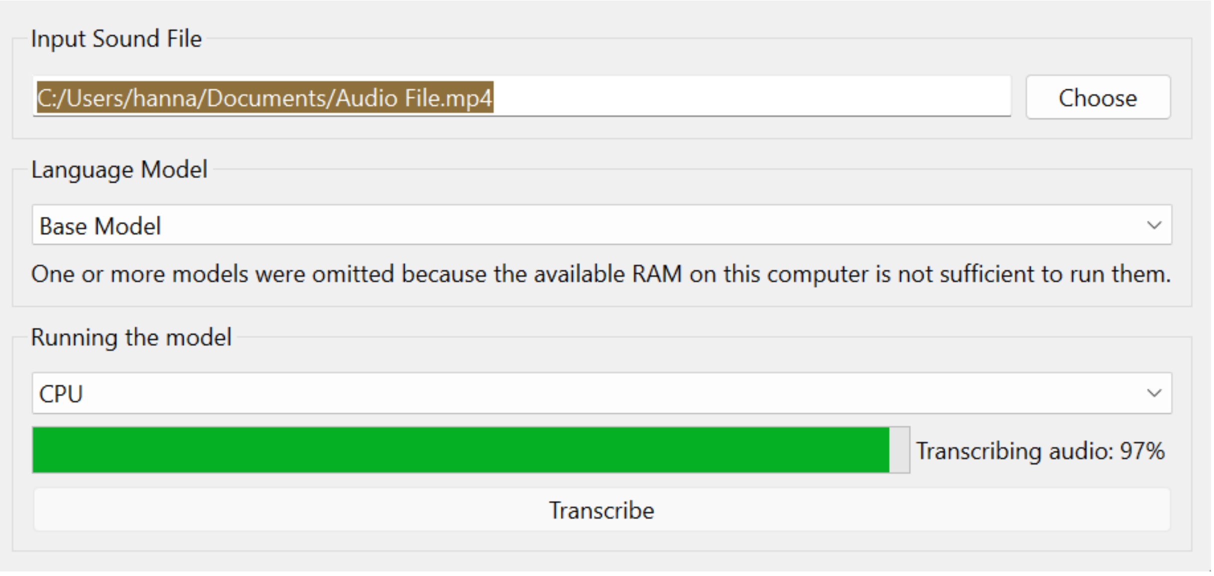


*The program needs to load the chosen language model. No data will be uploaded during the transcription process.*

**Once the transcription is finished a new pop-up window will appear with the generated text.**

You can now copy and paste the text into a document or program of your choice.
